# A qualitative study of music-based intervention use for Alzheimer’s disease in elder care communities

**DOI:** 10.1101/2025.03.18.25324196

**Authors:** Benjamin J. Hess, Ava Zatloukal, Jasmine M. Taylor, Michelle Neidens, Kristine N. Williams, Rebecca J. Lepping

**Author notes:** Corresponding Author: Rebecca J. Lepping, Ph.D., University of Kansas Medical Center, 3901 Rainbow Blvd, MS 2012 Kansas City, KS 66160.

## Abstract

**Background:** Because Music Based Interventions (MBIs) are not standard of care for Alzheimer’s disease and Alzheimer’s disease related dementias (AD/ADRD), it is likely that the application of them in different care communities differs widely. Additionally, there is no standardized use of personalized music listening and group music activities.

**Objective:** The purpose of this study was to assess the current use of music in long-term care communities, to identify trends and patterns of music use and record the observed benefits that music use provides.

**Methods:** This study utilized a qualitative research approach using semi-structured interviews with care community staff and care community observations to examine the role that music played as a therapeutic tool for individuals with AD/ADRD living in care communities.

**Results:** Five different communities were visited and observed. Of the five communities visited, interviews were conducted at four communities. One community did not participate in the interviews due to scheduling conflicts. Two staff members were interviewed at each participating community resulting in eight total interviews.

**Conclusions:** The results of this qualitative survey of care communities suggests that staff members believe that music use has beneficial effects for residents living with AD/ADRD. Music is economical, easily accessible, and very adaptable. Music can be used in a broad range of situations to improve the quality of life for both residents and staff in care communities. Music use can be active or passive, it can be used by an individual or a group to excite and engage or to calm and soothe.

---

Alzheimer’s disease and related dementias (AD/ADRD) are associated with a constellation of symptoms that can negatively impact quality of life. ^1–6^ Beyond memory loss and cognitive difficulties, AD/ADRD can worsen emotional health, impair functional capacity for activities of daily living, and block communication ability leading to social isolation. Music-based interventions (MBIs) are used to support social, emotional, and cognitive domains, with the goal of improving quality of life for people living with AD/ADRD and their families and caregivers. ^7–16^ Because MBIs are not standard of care for AD/ADRD, it is likely that the application of them in different care settings differs widely. Additionally, there is no standardized use of personalized music listening and group music activities. While music therapists are a part of some care teams, this is also inconsistent and limited for those in rural communities, leaving activity directors or other members of the care team to provide any MBIs or activities. The ambiguity caused by the potential variability in music use between care communities is something which needs to be clarified. This clarification can be accomplished by using qualitative semi-structured interviews with activity directors and music therapists and other care community staff. The purpose of this study was to assess the current use of music in long-term care communities, to identify standard trends and patterns of music use and record the observed benefits that music use provides.

## Methods And Procedures

### Research Design

This study utilized a content analysis research approach with semi-structured interviews and community observations to examine the role that music played as a therapeutic tool for individuals with AD/ADRD living in care communities. The objective was to discover different perspectives of memory care workers related to music use for individuals under their care. Semi-structured interviews were conducted with memory care workers to gather unique insights on the impact of music on everyday lives and the general wellbeing of community residents. The aim of this study was to identify key patterns and ideas centered around the effects that emerge specifically from the application of music in everyday settings within the care community. By focusing on the staff directly providing care, these interviews could reveal hidden insights regarding social interactions and emotional states that could not have been otherwise uncovered.

### Participants

The University of Kansas Medical Center Institutional Review Board (IRB STUDY00150618) provided ethical approval for this study. The study team obtained letters of permission to visit five care communities in the metro area of a large Midwestern US city. During these visits, care community staff were invited to participate in the study. Participants were provided with sufficient information regarding the purpose of the study and their confidentiality was ensured. Informed consent was obtained from all the participants before their participation, and participants were compensated for their time. Participants were selected based on their willingness to participate and their involvement in caring for residents with AD/ADRD. The inclusion criteria were activity directors or other staff engaged in MBIs at long-term care communities that are willing to be observed during MBIs and participate in semi-structured interviews. The exclusion criteria were activity directors or other staff engaged in MBIs at long-term care communities that are unwilling to be observed at MBIs or participate in semi-structured interviews. No specific age range was required.

### Study Sites

Care communities surrounding a large Midwestern US city were selected as potential study sites based on a set of predetermined criteria: whether the site was in an urban or rural area, the number of residential units and capacity of each community, the type of living arrangements offered (assisted living, independent living, etc.), and whether the community was one independent location or had multiple communities at different locations. These criteria were selected to ensure the selected sites reflected a wide range of community, living styles, degree of organization, and staffing levels. The selection process attempted to incorporate a wide representation of care communities with variation in size and number of residents. The potential care facilities also differed in their access to professional music therapists and the frequency of music related activities. This process allowed for a more complex understanding of how the different music usage affected individual’s outcomes.

### Data Collection Methods

Data collection consisted of semi-structured in-person interviews. During each interview, one researcher performed the role of “interviewer,” while a second researcher had the role of “recorder.” The interviewer went through a predetermined set of 20 questions to capture the participants’ view on music and how it affects residents with AD/ADRD (Supplemental Table 1). Throughout the interview process, follow-up questions were used to clarify or request expansion for participants responses, allowing for more individuality and unique perceptions. The conducted interviews were audio- and video-recorded via an iPad and were transcribed to a Microsoft Word document and uploaded for analysis into Dedoose Version 9.2.4. ^17^ Both the interviewer and recorder also took notes by hand during the interview process as a backup to the audio/video recordings.

### Observations

Observations were conducted by the research team in each of the care communities. The care community staff provided a tour of the shared spaces. During this tour, the environment, music, setting, activities, and interactions of the individuals were noted and recorded on a paper notepad that was later transcribed into a Microsoft Word document. The research team also documented anything related to music such as the presence and use of instruments, smart speakers, special equipment, record players, etc. When available, artifacts such as daily activity calendars were also collected. These observations were uploaded into Dedoose and compared alongside the observations and staff interviews of other care communities within the area.

### Data Analysis

Data analysis and structuring for this paper were based on the “Standards for Reporting Qualitative Research” checklist, which was created by O’Brien and colleagues, the EQUATOR Network was used to identify the most appropriate checklist ^18, 19^. Thematic analysis was used to analyze the data by identifying recurring themes that were identified in the text ^20^. This analysis process included reading through the transcriptions and first finding broad categories that related to the research. Afterwards, sub-categories were derived from overarching ideas, then general themes were recorded based on what was discussed during the interviews.

The transcription process consisted of taking the recorded data from each interview and typing in a Word document alongside the audio file. Transcription was conducted by one research staff member (AZ), and questions were verified with a second who was present during the interview (BH). Ambiguous sections of the audio recordings were compared with written notes taken during the interview. Once the first transcription was finished, the audio was reviewed a second time as the document was proofread and any errors were corrected.

After this process, the transcript data were coded into categories and sub-categories using an open coding method within the software Dedoose, a web-based software for managing and analyzing mixed methods research data, as described in *Qualitative Research and Evaluation Methods* ^21^. These codes were created based on concepts that were consistently brought up and mentioned across the eight different interviews. Following open coding, a round of axial coding began. Distinctive and subclusters were identified within the overarching categories and related to other quotes that shared the same theme in an iterative process ^20^ . Coding was conducted by two members of the study team (AZ, BH). Connections between these different identified themes were identified and described. The analysis process attempted to reveal the experiences of care community workers with music especially as it related to AD/ADRD care. To ensure credibility, peer debriefing was conducted to develop a more well-rounded understanding of the content.

## Results

The primary aim of this study was to explore the different effects of music on individuals with AD/ADRD through interviews conducted with caregivers and music therapists. The data were categorized into four main topics based on the predetermined interview questions: *Music Use, Music Effect, Level of Involvement,* and *Music for Care*. Through two rounds of coding (open and axial) and thematic analysis they were further subdivided to capture key ideas that emerged from the data. The findings showed different patterns in music influencing memory, mood, and the quality of care provided by the responses of each of the individuals interviewed.

### Site Characteristics and Interviewee Demographics

Ten care communities were identified as meeting criteria for this study. After contacting the ten care communities, five care communities agreed to participate in the study and were visited in person by the study team for observation and staff interviews. The communities ranged from 15 rooms to 176 rooms. The levels of care offered by the communities varied, overall levels of care included assisted living, independent living, rehabilitation, skilled nursing, long term care and memory care. All the participating sites were based in and around a large Midwestern US city.

Of the five sites visited, interviews were conducted at four sites. One site did not participate in the interviews due to scheduling conflicts. Eight individual staff members at these care communities participated in the interviews. Two staff members within four of the care communities, all of whom worked in direct contact with the residents, individually participated in a semi-structured interview, which resulted in a total of eight interviews. Interviewed staff had varying credentials and certifications and had been working in the communities anywhere from less than 1 year up to 15 years. The individuals that were interviewed had a variety of musical backgrounds ranging from no musical experience to having taken piano lessons to having musical degrees or being registered music therapists. Six out of the eight interviewees indicated they were highly involved in activity planning for their care community and served as program managers and activity directors. The other two interviewees were a medical aide and a music therapist. There were other roles represented, such as house manager and directors of sales engagement that also participated heavily in the activity planning process. Interviews were conducted on site at each community in a private room with a closed door. The interviews lasted an average of 27 minutes and ranged from 18-41 minutes.

### Coding

The codebook is included as Supplemental Table 2. Supporting quotes are numbered to differentiate between interviews.

#### Music Use

The first Dedoose coding category that emerged was *Music Use*, which highlighted the interview sections related to the music type that was being played, the occasion it was used, and how it was used with the individuals. The analysis revealed four subcategories: *Active Listening* (1a), *Passive Listening* (1b), *Type of Music* (1c) and *Frequency* (1d).

*Active Listening* was frequently mentioned in the interviews when the caregivers described individuals actively engaging with the music through different musical sessions by singing along, answering questions about the songs, or using instruments to create music. In these sessions, the individuals in the care community were encouraged to participate. These sessions were often described as being helpful for promoting engagement and stimulating memory.

“this crowd really loves singing so if one starts humming a tune they’ll all kind of just pick it up together and now they’re singing the lyrics as much as they do remember”-R5

“some people do like to just sit and listen and sing along and and um just enjoy it other people do like um shaking a tambourine or what have you and they get very serious about it”-R1

“she came and played and they sing along and sometimes she’ll pick out she’ll say what’s your favorite song and ask them and and they’ll have a response they’ll know their favorite song”-R2

“sometimes when we gather residents in memory care it’s like herding cats because you bring them there and they leave as you’re gathering more people but um the music really helps because people are like “oh there’s music and I know this song” um and then it energizes the residents and keeps them moving throughout the exercise itself”-R3

*Passive Listening* referred to times when music was played in the background to create a calm environment without active music participation. Interviewees described music as being used passively at different times throughout the day to calm or engage. These times included meals, activities, daily routines, and rest.

“best practice we found for music in the dining room is that if you are in the middle of a meal we tend to use non lyrical instrumental music […] what we find is if I put on Patsy Cline sometimes that resident that maybe needs to focus in on what they’re eating is going to be distracted by what Patsy’s singing and so having something that kind of stimulates but isn’t distracting that’s there’s a fine line there”-R4

“there’s always music playing in the background during breakfast lunch dinner time”-R2

“there might be somebody who doesn’t want to participate in in an activity but we still want them to be present and um be social and be engaged that way um with music in the background um oftentimes they end up just kind of being drawn into the activity and just um enjoying it just as much”-R1

“the minute I get here I have Alexa playing music and it pretty much goes all, all day and all evening”-R6

*Type of Music* was further subdivided into: Live Music, Instruments, Piano Playing, Smart Speakers, and Recorded Music. Live music was very common, and musicians frequently came into the communities to play. Instruments were sometimes provided to residents who wanted to participate in the live music. Piano playing was regarded as being a therapeutic form of music that could help individuals with AD recall different memories. There was frequent usage of smart speakers, with each of the communities having a minimum of one smart speaker, while some communities had smart speakers in multiple rooms. These devices were used to play recorded music that appealed to each of the residents’ preferences and could be easily adjusted to each individual or group. Recorded music, which was often preferred music from an individual’s past, was usually delivered using phones or smart speakers

“we also have someone come that passes out like the instruments that we had and they get them involved in doing music um in the morning time”-R2

“for Alexa I’ll go around and ask them “hey you know what’s one of your favorite songs?” and then I’ve kind of like gathered you know what they like and play what’s more appropriate for them”-R6

“the gentleman that comes with the guitar he comes twice a week to our unit, um he comes to other units, but to us he comes uh not twice a week, twice a month”-R9

“…we have live music on a rotation of different performers every week, all of which touch back into a lot of that older music, most of them will sing along to it” -R5

“we have the Alexas in all of our neighborhoods here in memory support”-R5

“Um, the way we involve them, we just I mean like I said she’ll have somebody come and we just gather them, I play music like the minute I get here I have Alexa playing music and it pretty much goes all, all day and all evening.” -R6

*Frequency* was created to note how much each of the care communities played music on an average day. *Frequency* was categorized into Time When Played which was further divided into Midday, Morning, and Night. *Frequency* varied depending on the location, average use was high according to the staff at the sites.

“Every day every hour every minute and somewhere around in this building if you walk around it you’ll be hearing music”-R8

“I mean throughout the 24 hours of a day um wow I haven’t thought about it in that way I guess I would maybe average it out to be probably 40 to 60% of the day probably 50 cause you, you don’t listen to it at night when you’re sleeping”-R3

“… music’s played quite a bit I’ve never actually it’s it’s making me think I should really think about like how much music is played here um but it’s quite a bit um I would say probably um you know 10 hours out of the day there is some form of music”-R1

“Daily, of the Alexas alone almost never turn off they’re on from the time they come in for breakfast in the morning till they go to bed at night.”-R5

#### Music Effect

The second Dedoose coding category, *Music Effect*, focused on the impact of music on memory restoration and mood alteration. It was divided into two subcategories *Memory Restoration* (2a) and *Mood Effect* (2b).

*Memory Restoration* was a frequently discussed subcategory of music effect. Participants often mentioned that the familiar music triggered the memory of past events and places. Different music from residents’ youth and significant life events stimulated memories and offered a sense of identity.

“…somebody remembers a song or somebody oh my brother loved that song and um you know my brother loved Johnny Cash or my dad loved Johnny Cash so it just it just you know kind of takes on a life of its own”. -R1

“then it sparks it to them and they’re singing the words and you know sometimes it’s it brings us to tears because maybe they haven’t even said a word or you haven’t heard them say much in a long time but they’re singing this whole song”-R9

“they don’t you know remember what we did 5 minutes ago but that song comes flooding right back to em and they can sing all the words perfectly, so it’s a great reminiscing aid”-R5

“I think it’s very impactful because something about the brain music is remembered even in the latest stages of Alzheimer’s and even if you lose the ability to speak you still might be able to hum your favorite song”-R3

“you know we’ve even gone and done a nursery rhyme you know and what they start to think about either from them being a child or having children um you know somebody always speaks out they’re like oh I always sing this song to my kids and things like that”- R1

*Mood Effect* was also frequently discussed. Effects varied from music relaxing and calming the individuals to energizing and engaging them. Music was reported as having many positive influences on emotional wellbeing. Different interviewees observed that the music alleviated the symptoms of anxiety and allowed for a calm and relaxed environment.

“kind of relaxes them either puts em to sleep or just kind of slows em down, uh calms that anxiety down”-R9

“today we had two women starting to argue um and so we turned on Sweet Caroline cause they love that song and so they, they instantly started singing and forgot that they were arguing. So I, I think it’s, it calms them down”-R6

“it can be used to be energizing and help bring people into a more positive mood and have more energy like when we do exercise and we use music in the background”-R3

“sometimes we’ll individualize and have them have their radio and place it right next to them which is a soothing calming thing for them as well”-R2

#### Level of Involvement

The third category *Level of Involvement* explored music interacting with activity planning and resident care methods. It was subdivided into *Activity Involvement* (3a) and *General Care* (3b).

*Activity Involvement* focused on music playing a role in activating the resident engagement in different activities. This was through musical games, dance, simple percussion instruments and other musical activities. This was further subcategorized into Musical Activity for activities that were specifically music focused and Non-Musical Activity for activities that were not music focused but were sometimes supplemented with music. Musical activity was considered effective in promoting physical and mental engagement amongst the individuals through instrument use and singing along. Participation or engagement was deemed essential for maintaining function and well-being.

“I always get good participation for music, music is something I can get more people out of their rooms for. Also, if they want to be a passive participant it’s nice as well, cause they can just listen and enjoy and don’t feel the need to have to participate in the group if they don’t want to, just to hang out with us, I’ll take it.” -R5

“we play music bingo we play um musical chairs we play cakewalk which it has music you know we have cards playin’ we’re we’re playing cards and we have music in the background”-R8

“I was thinking you know even different people who come in and do therapies and stuff um will have started to use music as well with our residents so um it’s just a nice addition to just you know you can work out and exercise or you can beat a drum to fun music and call it a workout”-R1

*General Care* emphasized the role of music in overall caregiving. Many caregivers mentioned that music played a role in calming an individual. Many caregivers also mentioned various tasks that they performed to assist in the resident’s overall day to day living that did not require any use of music and it was placed under this category.

“I get here, um we do, we get a report, I wake people up turn on lights, wake people up help em get dressed um serve them their breakfast, their coffee, um try to keep them occupied as much as possible with little things you know, if there’s nothing major going on then I will print um coloring papers off and so we’ll sit around the table and we color, we, we tal… we’ll, we’ll have coffee, we’ll have a group and we’ll have coffee or tea or whatever and we’ll talk about, um I’ll get on my phone and look up what happened on this day in history, so we’ll all talk about that, um we’ve sat around and talked about the weirdest foods we’ve had and they really enjoyed talkin’ about that.”-R6

“You know I as soon as you come in you’re you’re talking to them getting them up like singing them in the morning to get em in a better mood to wake em up you know different things like that helping them brush their teeth and comb their hair and hey let’s read this book or you know I think that’s probably 90% of all of my job here at ***** no matter what position that you have here you are actively engaging with the residents the whole time you’re here so you may have a little break to eat a sandwich but but you’re actively engaged with the residents here”-R2

#### Music for Care

The fourth category *Music for Care* regarded aspects of care that affected the overall quality of individual lives. They encompassed the impact of music on individuals’ wellbeing and improving the emotional, psychological, and social states of life for individuals with AD. Caregivers described the music as providing significant benefit to the daily lives of individuals, as well as being easy to access. The consistent exposure to music contributed to a more positive and stable environment.

“it’s a very easy thing to do it doesn’t cost any money especially now with all the streaming”-R1

“Activities provides this outlet where people can come in and they’re not worried about they’re not taking medications they’re not bein… having wound care done they are focused and they they have a chance to focus on themselves and say I’m gonna do exercise because that’s something that I do for self-care or I’m you know we’re gonna go play trivia or we have a storyteller that’s coming in I love Johnny Cash he’s gonna talk about Johnny Cash and for a minute there we’re not worried about where we are or who we’re around or where our family is we have a chance to kind of just be ourselves in that moment and I think that is a an incredibly important part of not just rehab but personal mental health and wellness”-R4

“Um we do live entertainment a couple times a week as more of a social experience we’ve experimented with a choir here to have that experience of singing as a group together […]and also to help with the diaphragmatic breathing for the rehab residents or some of the long term care residents who just need that support with their breathing overall and also with residents with Parkinson’s”-R3

## Discussion

Several recurring themes emerged throughout the interviews. Music was seen as easily accessible through phones and smart speakers which allows it to be used by all staff and by residents themselves. Music was seen to play a role in mood regulation for residents of the care communities living with Alzheimer’s disease. Caregivers used music to reduce stress and bring joy, lifting spirits and providing comfort to the residents. Music was also seen to be beneficial both as the primary focus and as a supplement to other non-musical activities. The results suggest that music played a useful role in the care of individuals with AD/ADRD through the usage of music with varying levels of involvement throughout each day. Music was used to calm as well as engage and was used both passively and actively. Music was seen to cause positive responses in typically unresponsive residents, and promote memory recall and sense of self, it also was seen to lessen anxiety. Caregivers can stimulate memory, positive mood, and can foster wellbeing by using music. Music was also described as facilitating a better caregiving process and aiding in emotional support for care community members. Some limitations to this study were a small sample size and all care communities being in the same geographic region of the Midwestern United States. There was also selection bias since all communities that agreed to participate used music as a part of their care community. Future research with a larger sample size and a wider range of locations would be needed to verify the results of this study. Additionally, research on the effectiveness of specific MBIs will help to define evidence-based best practices and guidelines for the use of music in elder care communities.

Music was generally viewed as beneficial by all the interviewed participants, although usage varied between care communities and ranged from generalized music listening to more structured music-based interventions. This seems to indicate that, regardless of the level of focused musical activity, music was seen to improve quality of life for residents with AD/ADRD. Even within each individual community, music was incorporated with various levels of focus, sometimes functioning as a background mood enhancer and sometimes as the primary focus of a sing-along or musical event. The adaptability of music for use in many situations makes it useful throughout most of the day across a wide range staff duties and resident needs. The ease of access to music, provided through smartphones and tablets, makes it an effective tool for staff in a variety of positions even if they are not an activity coordinator or a musical interventionist. Engagement in non-musical activities, such as exercise or physical rehabilitation, was also seen to be improved through the inclusion of music. This improvement seems to indicate that even when the music itself is not the focus of an activity, it can improve the appeal and the success of an activity by increasing participation and engagement. As stated in a quote from one of the interviewees, regarding the effect of music on individuals in general, “I just think music it’s very vital in life for everybody” -R2.

## Conclusion

The results of this qualitative study of care communities seem to show that music use is believed by care community staff to have beneficial effects for their residents with AD/ADRD. Music is economical, easily accessible, and very adaptable. Music can be used in a broad range of situations to improve the quality of life for both residents and staff in care communities. Music use can be active or passive, it can be used by an individual or a group to excite and engage or to calm and soothe. Music use has very few adverse effects. Given the positive effects that music has been observed to provide and the ease with which it can be used, further investigation must be done to better understand the neural and emotional mechanisms which drive these effects in individuals with AD/ADRD.

## Ethical Consideration

Ethical approval for this study was obtained from the KUMC Institutional Review Board (STUDY_00150618). Participants were provided with sufficient information regarding the purpose of the study and their confidentiality was ensured. Written informed consent was obtained from all the participants before their participation. All data were stored securely in databases following ethical guidelines.

## Data Availability

All data produced in the present study are available upon reasonable request to the authors

## Acknowledgements

We thank the staff and care community members who have shared their knowledge and enthusiasm for music as a support for people living with dementia. Research reported in this publication was supported by the National Center for Advancing Translational Sciences of the National Institutes of Health under the Award Number UL1TR002366 to Rebecca Lepping. The content is solely the responsibility of the authors and does not necessarily represent the official views of the National Institutes of Health.

## References

1. Patton MQ and Patton MQ. *Qualitative research and evaluation methods*. 3 ed. Thousand Oaks, Calif.: Sage Publications, 2002, p.xxiv, 598, 565 p.

2. Heru AM, Ryan CE and Iqbal A. Family functioning in the caregivers of patients with dementia. Int J Geriatr Psychiatry 2004; 19: 533–537. DOI: 10.1002/gps.1119.

3. Jutkowitz E, Brasure M, Fuchs E, et al. Care-Delivery Interventions to Manage Agitation and Aggression in Dementia Nursing Home and Assisted Living Residents: A Systematic Review and Meta-analysis. J Am Geriatr Soc 2016; 64: 477–488. DOI: 10.1111/jgs.13936.

4. Kales HC, Gitlin LN and Lyketsos CG. Assessment and management of behavioral and psychological symptoms of dementia. BMJ 2015; 350: h369. 20150302. DOI: 10.1136/bmj.h369.

5. Regier NG and Gitlin LN. Dementia-related restlessness: relationship to characteristics of persons with dementia and family caregivers. Int J Geriatr Psychiatry 2018; 33: 185–192. 20170323. DOI: 10.1002/gps.4705.

6. Regier NG, Hodgson NA and Gitlin LN. Neuropsychiatric symptom profiles of community-dwelling persons living with dementia: Factor structures revisited. Int J Geriatr Psychiatry 2020; 35: 1009–1020. 20200526. DOI: 10.1002/gps.5323.

7. Guetin S, Portet F, Picot MC, et al. Effect of music therapy on anxiety and depression in patients with Alzheimer’s type dementia: randomised, controlled study. Dement Geriatr Cogn Disord 2009; 28: 36–46. 20090723. DOI: 10.1159/000229024.

8. Hofbauer LM, Ross SD and Rodriguez FS. Music-based interventions for community-dwelling people with dementia: A systematic review. Health Soc Care Community 2022; 30: 2186–2201. 20220630. DOI: 10.1111/hsc.13895.

9. Jordan C, Lawlor B and Loughrey D. A systematic review of music interventions for the cognitive and behavioural symptoms of mild cognitive impairment (non-dementia). J Psychiatr Res 2022; 151: 382–390. 20220425. DOI: 10.1016/j.jpsychires.2022.04.028.

10. McCrary JM, Altenmuller E, Kretschmer C, et al. Association of Music Interventions With Health-Related Quality of Life: A Systematic Review and Meta-analysis. JAMA Netw Open 2022; 5: e223236. 20220301. DOI: 10.1001/jamanetworkopen.2022.3236.

11. Petrovsky DV, Gooneratne NS, Bradt J, et al. Tailored music listening intervention to reduce sleep disturbances in older adults with dementia: Research protocol. Res Nurs Health 2020; 43: 557–567. 20201102. DOI: 10.1002/nur.22081.

12. Román-Caballero R, Arnedo M, Triviño M, et al. Musical practice as an enhancer of cognitive function in healthy aging - A systematic review and meta-analysis. Plos One 2018; 13. DOI: ARTN e0207957 10.1371/journal.pone.0207957.

13. Särkämo T, Laitinen S, Numminen A, et al. Clinical and Demographic Factors Associated with the Cognitive and Emotional Efficacy of Regular Musical Activities in Dementia. J Alzheimers Dis 2016; 49: 767–781. DOI: 10.3233/Jad-150453.

14. Särkämö T, Laitinen S, Numminen A, et al. Pattern of Emotional Benefits Induced by Regular Singing and Music Listening in Dementia. Journal of the American Geriatrics Society 2016; 64: 439–440. DOI: 10.1111/jgs.13963.

15. Särkämö T, Tervaniemi M, Laitinen S, et al. Cognitive, Emotional, and Social Benefits of Regular Musical Activities in Early Dementia: Randomized Controlled Study. Gerontologist 2014; 54: 634–650. DOI: 10.1093/geront/gnt100.

16. van der Steen JT, Smaling HJA, van der Wouden JC, et al. Music-based therapeutic interventions for people with dementia. Cochrane Db Syst Rev 2018. DOI: ARTN CD003477 10.1002/14651858.CD003477.pub4.

17. Dedoose: cloud application for managing, analyzing, and presenting qualitative and mixed method research data. Version 9.2.4 [software]. SocioCultural Reseacrh Consultants, LLC. [cited 2025 Mar 18]. Available from: www.dedoose.com.

18. O’Brien BC, Harris IB, Beckman TJ, et al. Standards for reporting qualitative research: a synthesis of recommendations. Acad Med 2014; 89: 1245–1251. DOI: 10.1097/ACM.0000000000000388.

19. equator-network.org [Internet]. EQUATOR Network | Enhancing the QUAlity and Transparency Of health Research [cited 2025 Mar 18]. Available from: https://www.equator-network.org.

20. Marshall C and Rossman GB. Designing qualitative research. Sixth edition. ed. Los Angeles, California: SAGE, 2016, p.xxii, 323 pages.

21. Patton MQ. Qualitative research & evaluation methods : integrating theory and practice. Fourth edition. ed. Thousand Oaks, California: SAGE Publications, Inc., 2015, p.xxi, 806 pages.

