## Supplemental Information for "A qualitative study of music-based intervention use for Alzheimer’s disease in elder care communities"

**Supplemental Table 1: Interview Questions**

To get us started, I would love to get your thoughts and perspective on Alzheimer’s Disease and the role music can play in caring for individuals living with Alzheimer’s Disease and related dementias.

1) How do you think musical activities could be used in caring for people living with AD/ADRD?

2) How would you describe the impact of music for residents living with AD/ADRD?

3) Are there specific aspects of care that would be more impacted by musical activities?

I want to shift the conversation just a bit and talk further about the musical activities your care community provides to get a general idea of the unique atmosphere of the care community and your selection of music.

4) Do any of the activities for the residents involve music and if so, can you describe the ways the care community involves music in everyday activities? (listening, playlists, singing, music therapy, etc.)

5) ***can ask 5-7 if they play music over an intercom/throughout community*** How is music usually selected? (is there a music selection process, is it selected by one person or chosen by a committee or staff members?)

6) When playing music, is your care community consistent and stick to one song/playlist or is there more variety with song changes (different rhythms, tempos, etc.)

7) How often is music played? Is music only played during certain times or is it constantly played and rarely turned off?

8) How much do the care team, residents, or residents’ family members’ opinion contribute to the various activities the resident participates in?

9) How often is music a part of activities?

10) Do you think the current amount of music is sufficient?

11) Has the use of music activities changed in this care facility over time? If so, in what ways?

Now that we’ve talked more in-depth about the ways your community incorporates music into daily living activities and the atmosphere it provides, I would love to talk further about your position and how your role plays a part in the care community.

12) What is your role in this care community?

13) What would a typical day in your position look like?

14) How are you actively involved in caring for people living with Alzheimer’s disease or related dementias specifically?

15) How much do you participate or provide in activities with residents?

16) How involved are you in creating activities or activity planning?

17) What is your background or experience with music? For example, do you have any formal training?

18) Finally, do you have anything you would like to add that we did not cover?

Thank you for your time, your answers and time are greatly appreciated.

**Supplemental Table 2: Dedoose Codebook**

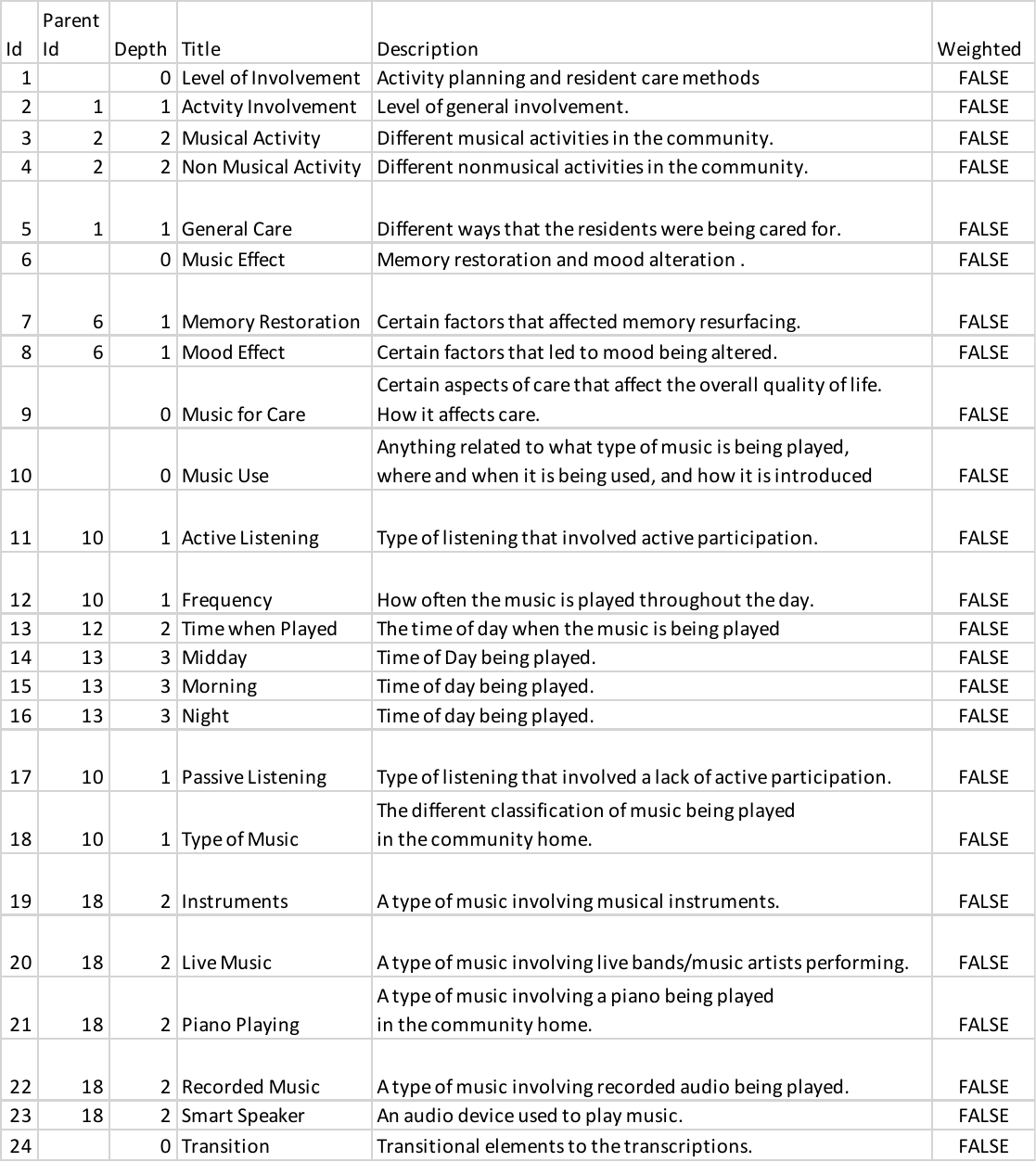
